# Co-Designing HPV Vaccine Programs with Girls and Caregivers: Insights from a Human-Centered Design Approach

**DOI:** 10.64898/2025.12.16.25342426

**Authors:** Eliza Fishman, Nicole Castle, Munyaradzi Joel Chinguwa, Amenze Eguavoen, Temitope Adebola Alfred, Steven Ellis, Yeni Indra, Paul Bassi Amos, Brittany Iskarpatyoti

**Affiliations:** JSI Research & Training Institute, Arlington, Virginia, United States of America; JSI Research & Training Institute, Harare, Zimbabwe; Lluvia Health Organization, Lagos, Nigeria; Independent Consultant, Abuja, Nigeria; Empatika, Jakarta, Indonesia; JSI Research & Training Institute, Abuja, Nigeria; JSI Research & Training Institute, North Carolina, United States of America

## Abstract

Cervical cancer is a significant global health issue, ranking as the fourth highest cause of cancer-related deaths among women globally. HPV vaccination for girls aged 9–14 offers a powerful tool for cervical cancer prevention, yet global coverage remains low, particularly in low- and middle-income countries. While research has documented parental and stakeholder perspectives, few studies center on girls’ own experiences and how gender norms shape their access and decision-making. This study applied a human-centered design (HCD) approach in Nigeria and Indonesia to explore barriers to HPV vaccination and co-develop context-specific solutions with girls, caregivers, and immunization stakeholders.

Eight co-creation workshops were conducted with adolescent girls (ages 10–17) and caregivers in Nigeria (n=64) and Indonesia (n=66). Using HCD techniques—including persona development, journey mapping, and prototyping—participants identified barriers, enablers, and potential solutions. Stakeholder workshops in both countries validated and refined these community-generated ideas. Data were analyzed thematically using UNICEF’s Journey to Health and Immunization framework.

Across both countries, girls and caregivers reported limited awareness and inconsistent information about HPV vaccination. Misinformation, unclear communication, and insufficient preparation were common. Gender norms strongly shaped decision-making: while mothers typically led discussions, fathers or male community leaders often held final authority, particularly in Nigeria. Access challenges included transportation costs, distance to facilities, missed opportunities during school-based delivery, stockouts, and limited guidance on managing adverse events, with out-of-school girls experiencing the greatest barriers.

Co-designed solutions emphasized improving communication through trusted messengers, strengthening coordination between schools and health facilities, increasing engagement with fathers and community leaders, and expanding flexible, community-based delivery models to reach out-of-school girls. Findings underscore the importance of centering girls in HPV program design and demonstrate the value of HCD in generating actionable, locally relevant strategies.

Strengthening communication, addressing gender norms, and improving equitable service delivery are critical to increasing HPV vaccination uptake and advancing cervical cancer prevention.

## Introduction

Cervical cancer is a preventable and treatable disease, yet it ranks as the fourth highest cause of cancer-related deaths among women globally, disproportionately affecting women in low- and middle-income countries (LMICs) where access to primary and secondary prevention and treatment are constrained (1,2). In 2023, only 27 percent of girls globally received a single dose of HPV vaccine. There are large disparities in the distribution of national vaccination programs and vaccination coverage rates between high-income countries (HICs) and LMICs, with 88 percent of HICs having included HPV vaccination in their national immunization program, compared to 40 percent of LMICs (3,4).

There have been many challenges to introducing and increasing HPV vaccination coverage in LMICs, including supply and logistical constraints, COVID-19 pandemic disruptions, high vaccine costs, global and national funding for vaccination programs, vaccine prioritization, and health system capacity (1,4,5). Parents cited specific barriers to HPV vaccinations, such as cost, transport, and time constraints; low levels of knowledge about cervical cancer and HPV vaccination; and negative experiences with other vaccinations (6–11). Vaccine access and coverage are also challenged by misinformation, socioeconomic constraints, and gender inequities within and across countries. It is well known that gender and social norms affect decision-making, access, and uptake of HPV vaccination (12–15). These barriers are particularly pronounced in efforts to reach out-of-school girls, ensure completion of the vaccine series, accurately estimate target populations, monitor program performance, and sustain vaccination efforts (1,6). However, new opportunities are emerging to address these challenges, including the entry of additional vaccine manufacturers and the WHO recommendation for alternative single dose scheduling (16,17).

Evidence suggests that social norms related to peer approval and trust in the government and health professionals affect parental acceptance of and decision-making around HPV vaccination for their daughters (11,18). In many LMICs, mothers, typically the primary caregivers, often need to seek permission from their husbands to vaccinate their children. Women’s daily responsibilities, along with financial and mobility constraints, can further limit vaccine access and use (7,8,19).

While implementation strategies for HPV vaccination and studies on parental perspective have been documented, few studies explore the role of girls themselves in identifying key challenges or shaping programs designed to boost vaccine uptake (20). One study on adolescent involvement in HPV vaccine decision-making found that most had little to no influence, with over half remaining silent during the decision phase. This lack of influence was linked to their late or absent participation, underscoring the importance of engaging adolescents earlier in the decision-making process (21). Less is known about the challenges girls face and their priorities, as well as the social and gender-related factors that influence their uptake of the HPV vaccine. These factors are often overlooked in the design and implementation of vaccination programs. Engaging girls early in HPV program planning and decision-making may deepen our understanding of the factors influencing vaccination and help design solutions that strengthen and sustain high coverage.

## Country Context: Nigeria

In Nigeria, cervical cancer is the second most common cancer among women and the second most prevalent cause of cancer-related mortality among women aged 15–44 years (22). In 2022, there were more than 13,600 new cases of cervical cancer and more than 7,000 deaths from the disease (23). The Government of Nigeria introduced the HPV vaccine nationwide for girls aged 9–14 years in October 2023. The first phase of the introduction covered 16 states, and more than 4.8 million girls were vaccinated (24). In May 2024, the government implemented the second phase of the introduction for the remaining 21 states. As of December 2024, more than 12 million eligible girls have been vaccinated, bringing vaccination coverage to 71 percent (25). Following the campaign, Nigeria incorporated the HPV vaccine into its routine immunization schedule. The vaccine is delivered through school- and health facility-based programs. While the introduction campaign was successful, increasing vaccination coverage through the routine immunization system has its own set of challenges. To ensure program sustainability in Nigeria, a deeper understanding of the social and gender-related factors influencing HPV vaccine access and uptake from girls’ perspectives is important for designing tailored interventions that can improve and sustain high vaccination coverage nationwide.

## Country Context: Indonesia

Cervical cancer is the second most common cancer among women in Indonesia. In 2023, there were almost 37,000 new cases of cervical cancer, and more than 20,000 women died from the disease (26,27). In November of the same year, the president announced the National Plan for Cervical Cancer Elimination in Indonesia (2023–2030) (28). The Indonesian government introduced the HPV vaccine in Jakarta in 2016 to girls in school years five and six through a free, school-based program. Since its introduction in Jakarta, the government has been scaling vaccination nationwide through schools and health facilities; as of 2023, 132 municipalities are delivering the program to in-school and out-of-school girls aged 11 and 12 years (9,29,30).

According to the WHO, HPV vaccination coverage reached 95.9 percent for the final dose in 2023. However, it is unclear if that data includes out-of-school girls (31). Reducing barriers to vaccination and more effectively reaching out-of-school girls with solutions that center girls in program planning will be important to maintaining high levels of coverage as the Indonesian government expands the HPV vaccination program and advances its cervical cancer elimination goals.

## Study Objectives

Addressing the barriers girls and women face and increasing global HPV vaccination coverage requires the development and implementation of tailored, multi-level vaccination programs that meet the needs and preferences of girls and caregivers. This is true in both Indonesia and Nigeria as they integrate the HPV vaccine into their national immunization schedules. This study explores girls’ and their caregivers’ experiences, needs, and perspectives around improving HPV vaccination programs in both countries. The primary objective of this study was to explore both gender-related and non-gender-related barriers to the delivery and uptake of HPV immunization, while identifying potential solutions to address these barriers, as proposed by girls and women themselves and vaccine program stakeholders.

## Materials and Methods

### Study Design

This study employed a qualitative research design, using human-centered design (HCD) approaches to capture the experiences and perspectives of research participants. HCD is an iterative, solution-focused methodology that emphasizes understanding and prioritizing the needs, challenges, and experiences of end users and others directly involved in a system—in this case, girls, their caregivers, and vaccine program stakeholders (32,33). The study was approved by JSI Institutional Review Board (IRB) under the IRB reference (#24-37). The JSI IRB is registered with the Office of Human Research Protections (OHRP) as IRB00009069. JSI has a Federal-wide Assurance (FWA) on file [FWA00000218] and is a recognized IRB organization [IORG0007551]. Local ethics approval was received in both Indonesia and Nigeria. Written consent was obtained by all study participants.

### Study Population and Sample Selection

The study involved girls (ages 10–17) and their caregivers from four selected communities—two in Nigeria and two in Indonesia. Communities were purposively selected to capture high- and low-rates of out-of-school girls, urban/rural dispersion, and, where possible, religious diversity. Participants included equal distribution of vaccinated and unvaccinated girls, along with their caregivers. Caregivers were predominantly mothers; however, in both Indonesia and Nigeria, there were a few instances where non-maternal caregivers—such as grandmothers or female boarding school caretakers—were involved.

This purposive sampling approach enabled the collection of rich, relevant insights from those with direct experience of or influence over the vaccination process, including vaccinated and unvaccinated caregivers and girls, EPI managers, and representatives from implementing NGOs, multilateral agencies, and local partners. Results were shared with and refined by local vaccine program stakeholders, ensuring that the insights gained were actionable and relevant to program implementation.

### Sample Size

A total of 130 participants were included across eight workshops, with eight girls and their female caregivers (16 participants) in each workshop; one workshop in Indonesia included nine girls and their caregivers. Seventy stakeholders participated in the dissemination workshops held in Nigeria and Indonesia—45 in Nigeria (across two sub-national workshops) and 25 in Indonesia (in one national workshop). See Table 1 for sample characteristics.

**Table 1:** Sample Characteristics.

| Country | Nigeria |  |  |  | Indonesia |  |
| --- | --- | --- | --- | --- | --- | --- |
| States | Benue |  | Adamawa |  | East Java | West Java |
| Religion | Christianity |  | Islam |  | Islam | Islam |
| Study Site | Gwer-West | Makurdi | Fufore | Toungo | Bekasi City | Kediri Regency |

| Setting |  | Rural | Urban | Semi-urban | Rural | Urban |  | Rural |  |
| --- | --- | --- | --- | --- | --- | --- | --- | --- | --- |
| School Status Sampled |  | In-school | Out-of-school | In-school | Out-of-school | In-school | Out-of-school | In-school | Out-of-school |
| Number of girls sampled | Vaccinated | 4 | 4 | 4 | 4 | 5 | 5 | 4 | 3 |
|  | Unvaccinated | 4 | 4 | 4 | 4 | 3 | 4 | 4 | 5 |
| Number of caregivers sampled |  | 8 | 8 | 8 | 8 | 8 | 9 | 8 | 8 |
| Number of Stakeholders |  | 20 |  | 25 |  | 25 |  |  |  |

### Data Collection and Analysis

Data were collected at each site through two-day co-creation workshops, where participants—girls and caregivers—worked collaboratively to identify barriers to HPV vaccination and generate locally feasible solutions. Co-creation workshops are one method that can be used as part of the HCD process to conduct participatory activities to identify insights and generate solutions (34–36). During the workshops, participants developed personas, a tool used to create fictional characters that reflect user group characteristics and needs (32,37). Personas represented girls and their caregivers from their community who either had or had not been vaccinated.

Participants explored girls’ and caregivers’ characteristics, influences, experiences with health and HPV vaccination, values and aspirations, and gender and community norms. Following this, journey maps visually documented each persona’s experiences accessing and taking (or not) the HPV vaccine. Journey maps are used to understand key steps an end user or other stakeholder takes as they experience a product or service (32,38). UNICEF’s Journey to Health and Immunization was used as the journey map framework (39). It was developed to follow a caregiver’s journey to vaccinating a child while taking into consideration the health provider’s journey, and it has been used by other programs applying an HCD approach (35,36). It includes six domains that represent steps on the journey: 1) knowledge and awareness, 2) intent, 3) preparation, cost, and access, 4) point of service, 5) experience of care, and 6) after service (40). The journey maps supported caregivers and girls to identify key barriers to HPV vaccination based on their experiences at each step. After identifying key challenges, participants brainstormed ideas for potential solutions and developed prototypes, a version of an idea that can help communicate a concept and be tested (32). Following the community-level workshops, the personas, journey maps, and solutions were summarized and shared with relevant immunization stakeholders at the national (Indonesia) and sub-national (Nigeria) levels. A one-day workshop (two state-level workshops in Nigeria) was conducted to review outputs from community workshops, refine solutions, and develop action plans.

These interactive sessions served as the main method of data collection, capturing qualitative insights and actionable recommendations. Data were analyzed thematically based on the domains of the UNICEF Journey to Health and Immunization framework. The study lasted approximately 10 months, from June 2024 to April 2025, which included the planning and execution of workshops, data collection, and analysis. Recruitment began and ended at different times at each of the study sites. In Nigeria, data collection started in 01/09/2024 and ended in 28/02/2025. In Indonesia, data collection started in 01/10/2024 and ended in 09/04/2025.

## Results

The results are organized according to the domains of the UNICEF Journey to Health and Immunization framework, exploring the barriers and enablers girls and their caregivers encountered across each stage of the vaccination journey and follows with the solutions proposed by participants and reflected upon by stakeholders to address these challenges.

### Knowledge and awareness

Overall, in both countries and regardless of vaccination status, girls and their caregivers had varying but generally limited awareness and understanding of HPV vaccination. The ways they received information differed by country and vaccination status, but a similar pattern emerged: some unvaccinated caregivers in both Indonesia and Nigeria first heard about the vaccine through neighbors or social media.

### Indonesia

In Indonesia, vaccinated girls reported receiving information primarily from their mothers. Most mothers received their information from formal school communications delivered through letters, WhatsApp messages, or community sessions. While these communication mechanisms afforded more systematic outreach, participants expressed that information frequently arrived late or lacked sufficient detail to meet caregivers’ needs or support them to prepare their daughters for vaccination. Caregivers of vaccinated girls in Indonesia specifically ranked social media as the least trusted platform for HPV information, as it causes confusion and misunderstanding.

### Nigeria

In contrast, girls in Nigeria accessed information from a wider array of sources, including peers, radio, town announcers, and health workers. Many out-of-school girls reported being exposed to a variety of informal sources that often conveyed conflicting or inconsistent information.

Caregivers reported a similar pattern, drawing on diverse informal channels such as friends, daughters, religious leaders, and radio broadcasts. While this broad network enabled widespread dissemination, the compilation of information shared was described as often insufficient or unclear, limiting individuals’ ability to make informed decisions. As a result, caregivers’ access to accurate information varied widely, with some unaware of vaccination campaigns until after they had concluded. Girls also noted that women generally do not attend mosque and therefore miss out on important health messages that men receive there, as the messages are not further relayed.

### Intent and decision-making

Across both countries, female caregivers (often mothers) consistently confirmed their active role in decisions about HPV vaccination. Mothers were viewed as the primary caregiver responsible for their daughters’ health, and they typically guided vaccination choices. However, girls in both settings often had limited visibility into how decisions were made within the household, reflecting broader norms around obedience, modesty, and deference to parental authority.

### Indonesia

In Indonesia, mothers overwhelmingly described themselves as the main decision-makers regarding HPV vaccination, noting that fathers usually deferred to them. Girls also tended to rely almost exclusively on their mothers for guidance, with little mention of fathers’ involvement.

Despite their central role, many Indonesian mothers reported feeling under-informed and pressed for time when making vaccination decisions. They expressed concerns about misinformation and anti-vaccine narratives circulating in their communities and, in some cases, wished fathers were more engaged in discussions related to health-related decision-making processes.

### Nigeria

In Nigeria, mothers were also responsible for overseeing vaccination, but decision making was more collaborative than in Indonesia. Fathers and extended family members—particularly men—played a significant role, especially for unvaccinated girls. Girls noted that male approval was often required, and mothers frequently consulted husbands, relatives, community members, and religious leaders before deciding. Among caregivers of vaccinated girls, most sought information from health professionals and supported the vaccine, though several expressed frustrations with unclear or absent consent procedures. For unvaccinated girls, the need for male approval and financial support emerged as a key barrier, and women who made decisions independently were sometimes perceived as “rebellious” or faced social backlash.

### Preparation, cost, and access

Across Indonesia and Nigeria, both vaccinated and unvaccinated girls reported preparation, cost, and access as consistent barriers to HPV vaccination. Girls in both contexts described general challenges in accessing health services, including long distances to facilities, transportation-related costs, and limited availability of health workers. Caregivers in both countries echoed these issues, noting that insufficient information, unclear guidance, and logistical hurdles made it difficult for girls, particularly those out of school, to obtain the vaccine.

### Indonesia

In Indonesia, unvaccinated girls showed strong interest in receiving the HPV vaccine, partly because some believed it was free, though they were often unsure about the actual cost. Their primary barrier was a lack of information on how to access the vaccine—such as cost, eligibility requirements, or necessary documentation. While parental permission was a concern, it was secondary; some out-of-school girls even noted they might be able to persuade their parents to agree. For in-school girls, a major concern was not knowing where to go if they missed school-based vaccination sessions. Caregivers of in-school girls wanted greater involvement in the school vaccination process, citing insufficient notice and unclear instructions. Caregivers of out-of-school girls highlighted potential administrative barriers, such as the need for ID cards and interactions with unwelcoming facility staff.

### Nigeria

In Nigeria, girls—especially the vaccinated ones—identified barriers their peers might face, including long distances to schools where vaccination occurred and social pressures, such as losing friends who opposed the vaccine. Unvaccinated out-of-school girls emphasized two main obstacles: the distance to health facilities and the limited availability of parents, particularly mothers, to accompany them due to work, chores, fatigue, or travel. Some girls were also discouraged by peers and family members, including older sisters. For caregivers of both vaccinated and unvaccinated girls, transportation costs were a dominant barrier. Caregivers of vaccinated girls often accompanied their daughters but worried about lost income or business.

Those whose daughters missed the in-school campaign were directed to health facilities but encountered vague instructions, long travel distances, and inconsistent vaccine availability, frequently arriving to find the vaccine unavailable or the campaign already over.

### Point of service & Experience of Care

Across both Indonesia and Nigeria, girls generally viewed health workers positively, though their experiences varied widely depending on the setting and the individual provider. In both countries, girls and caregivers described logistical and communication challenges that affected the vaccination experience, such as limited information about the vaccine, insufficient guidance on managing potential adverse events, and procedural gaps during vaccination sessions. Participants in both countries also faced system constraints, including long wait times, crowding, and inconsistent availability of vaccines, which created barriers for girls and caregivers. For out-of-school girls in both Indonesia and Nigeria, accessing services at health facilities often involved delays, confusion, or stockouts, making the process more difficult than for in-school girls.

### Indonesia

In Indonesia, vaccination primarily occurred within school settings, which shaped most girls’ experiences. Girls reported a mix of positive and less positive interactions with health workers, but several noted feeling shy, uncomfortable, or excluded, especially those choosing not to get vaccinated. Some girls were separated from their peers during vaccination activities, which created confusion and social discomfort. Communication gaps were pronounced—many girls felt unable to ask questions or seek clarification during the session. The school-based model also limited access for out-of-school girls, who had to visit health facilities instead. These girls described long waits of up to an hour before receiving the vaccine. Caregivers in Indonesia generally understood the steps involved in school-based vaccination—registration, checks, and instructions for adverse event management—but felt that teachers and parents needed to provide more support. Some caregivers said teachers were unprepared to answer questions, and caregivers of unvaccinated girls who opted out reported being treated poorly during the process.

### Nigeria

In Nigeria, girls accessed vaccination both in schools and at health facilities, giving rise to more diverse experiences. While health workers were generally perceived positively, systemic issues, such as wait times and vaccine shortages, and poor crowd control were common. Some girls were unable to get vaccinated due to stockouts at health facilities while other in-school girls ran off during the campaign out of fear. Caregivers in Nigeria described generally positive vaccination experiences regardless of the delivery point. However, they noted that neither girls nor caregivers were adequately informed about potential adverse events or how to manage them. Echoing the girls’ accounts, caregivers highlighted logistical frustrations—including long delays, disrupted class schedules (for in-school girls), and the challenge of traveling to health facilities where vaccines were sometimes unavailable or health workers were unwelcoming (for out-of-school or unvaccinated girls).

### Post-vaccination experience

Post-vaccination experiences were shared only by vaccinated girls, as unvaccinated girls had little to report. Across both Indonesia and Nigeria, post-vaccination feedback from caregivers was generally positive, with caregivers pleased that their daughters were vaccinated and reporting few side effects. In both countries, neither girls nor caregivers described any severe adverse reactions, and overall satisfaction with having completed vaccination was high.

### Indonesia

In Indonesia, vaccinated girls reported minor side effects and noted a lack of guidance on post-vaccination care. Some caregivers, particularly in Kediri, shared perceptions that differed from the girls’ experiences; they were pleased with the vaccination outcome, while girls expressed frustration about returning to schoolwork immediately after vaccination and a lack of guidance on post-vaccination care.

#### Nigeria

In Nigeria, vaccinated girls generally expressed happiness and enthusiasm, often encouraging peers to get vaccinated. They reported no significant barriers or negative post-vaccination experiences. Caregivers in Nigeria did raise some concerns about side effects and limited information on post-vaccination care, but, like Indonesia, no severe adverse reactions were reported.

#### Proposed Solutions

Girls and caregivers developed solutions to address the challenges they had identified. State- and national-level stakeholders were engaged to reflect on the findings, share their own experiences, and collaboratively identify and prioritize actionable solutions. Each group’s solutions are summarized in Table 2 and Table 3.

**Table 2:** Summary of Solutions Developed by Girls, Caregivers, and Stakeholders in Nigeria.

| Subtheme | Girls | Caregivers | Stakeholders |
| --- | --- | --- | --- |
| Communication/Awareness | · School programs: Information should be delivered by the teachers or health educators during free periods and through peer-to-peer education to share information on vaccine safety, side effects, and benefits. Include motivators/incentives like provision of school supplies, etc.<br>· Community awareness: Messages should be shared through town announcers in the mornings and evenings. Comprehensive health education should be delivered by qualified health care personnel to ensure credibility and engagement. | · Use multiple communication channels including town announcers, community leaders, radio, tv, teachers, school authorities, religious houses, traditional leaders, women groups, Facebook, etc. to inform the public about the vaccines; if one channel doesn't serve them, another will.<br>· Vaccine champions should be identified in the communities to share their experiences and raise awareness about the vaccine in the community. | · Use multi-channel communication strategies, including town announcers, community leaders, and structured outreach programs to enhance knowledge and acceptance. |
| Coordination/Outreach | · School peer to peer education: train in-school girls to train or educate other out-of-school girls. | · Men should be engaged to obtain their support and buy-in as influencers and decision makers. | · Closer collaboration with local security forces and community networks can help ensure safe delivery.<br>· Increase targeted engagement with men. |
| Service Delivery | · Out-of-school peer-to-peer education enhances confidence and decision-making skills as well as increases girls' | · House-to-house vaccination should be considered because of its potential to reach |  |
|  | happiness. Peers can refer or accompany others to the health facility for vaccine uptake. | both in school and out-of-school girls. |  |
| System Strengthening/Supply Chain |  | · Improve school vaccination program by including proper planning to prevent stock-outs, more vaccinators, class-to-class vaccination instead of an assembly of all eligible girls, lesson free vaccination days where possible, a system to identify missed opportunities, and bidirectional communication with health care workers. | · Improve supply chain management and real-time tracking system. |

**Table 3:** Summary of Solutions Developed by Girls, Caregivers, and Stakeholders in Indonesia.

| Subtheme | Girls | Caregivers | Stakeholders |
| --- | --- | --- | --- |
| Communication/Awareness | · Display a poster with information on how to search for trusted information on HPV and HPV vaccination, including how to find information about HPV vaccination on TikTok.<br>· During in-school vaccination, use storytelling to pass the time and provide information about health and the vaccine for those waiting to get | · Organize educational activities at schools in coordination with puskesmas (public health clinics) to equip girls with information about HPV vaccination.<br>· Provide one-page leaflets from doctors/nurses for girls to show their parents.<br>· Provide socialization at schools two days before vaccination day to | · Develop engaging educational materials on HPV and the vaccine, including videos and interactive storybooks.<br>· Increase strategic collaboration with religious leaders to disseminate accurate information and address faith-based concerns. |
|  | vaccinated.<br>· Provide a fun, informative video before vaccination. | provide more information about HPV (with video and printed materials).<br>· Organize FAQ meetings for parents led by the health department. | · Create a comprehensive FAQ resource on HPV vaccination. |
| Service Delivery | · Provide education sessions at schools for girls and their parents that are facilitated by teachers and health workers.<br>· Organize outreach led by the puskesmas in coordination with the community and pesantren (Islamic boarding schools). | · Organize community socialization about HPV vaccination in communities residing around landfills.<br>· Teachers should include parents in WhatsApp groups to inform them of the vaccination process (at least three days before).<br>· Establish a coordination and collaboration process for schools and health centers, including a coordination system connecting the puskesmas with parents/caregivers. |  |
| System Strengthening/Supply Chain |  |  | · Implement standardized, child-friendly service delivery protocols. |

Girls in both countries suggested solutions centered on improving awareness and knowledge. In Indonesia, proposed strategies often focused on improving individual knowledge—such as making information clearer and more accessible for girls—but also reflected group-oriented elements, including peer support among girls and a desire among caregivers for community-based information sharing and collaboration with schools. Nigerian girls, on the other hand, predominantly recommended community-level interventions involving health workers, community leaders, and peers to improve vaccine uptake. Across both countries, girls emphasized the importance of timely, consistent information and improved access to vaccination services. Additional solutions focused on enhancing the vaccination experience and improving access.

Caregivers in Nigeria focused their suggestions on community-driven solutions, including using local leaders, town announcers, media campaigns, and home visits to raise awareness and combat misinformation. Incentives and logistical support (e.g., transportation vouchers or providing food during information sessions) were also recommended to improve access. Indonesian caregivers for in-school girls emphasized school-centered approaches and saw girls themselves as effective messengers in convincing parents about the value of vaccination. They also highlighted the need for better coordination among schools, teachers, and health workers, as well as improved communication to prepare families for vaccination. These proposed solutions closely aligned with those expressed by the girls, reinforcing the shared priorities of improved knowledge, support, and access.

Similarly, stakeholders in both Nigeria and Indonesia noted low public awareness and widespread misinformation as a critical challenge to their programming. In Nigeria, stakeholders emphasized the importance of multi-channel communication strategies, proposing the use of town announcers, community leaders, and structured outreach programs to enhance knowledge and acceptance. In Indonesia, communication between schools and parents is channeled through schools, but there is a lack of clear processes and standardized guidance on how schools should engage with caregivers, including what information to share, when to share it, and in what format. To address this, stakeholders proposed the development of educational and informational materials for both girls and their caregivers.

Stakeholders in both countries also focused heavily on logistical and programmatic operation challenges. Operational hurdles in Nigeria—such as vaccine stock-outs, delays in distribution, and incomplete data reporting—were seen as major impediments to effective rollout.

Stakeholders underscored the need for improved supply chain management and real-time tracking systems. They also advocated for closer collaboration with local security forces and community networks to ensure safe delivery in conflict-affected regions. In Indonesia, access-related issues were compounded by the lack of engagement with marginalized groups and data collection gaps, particularly for unregistered pesantren (Islamic boarding schools). At the point of service delivery, stakeholders noted the unclear delineation of responsibilities between schools and health services and weak cross-sectoral coordination.

Stakeholders in both Nigeria and Indonesia identified that greater engagement is needed with men, as fathers, decision makers, and religious and community leaders. However, stakeholders also stressed the importance of promoting gender inclusivity within HPV vaccination programs to ensure that strategies are responsive to the needs of girls and their caregivers.

## Discussion and recommendations

### Gender Norms and Decision-Making

The findings underscore the significant influence of gender norms on decision-making dynamics and access to HPV vaccination in both Indonesia and Nigeria. Immunization programs in LMICs have traditionally focused on women as the primary caregivers for children’s health (8,41).

However, existing literature also emphasizes men’s dominant role as household decision-makers (42–47). This study found this duality plays an important role in overall acceptance of the vaccine. While caregivers and girls in both contexts frequently identified mothers as the primary decision-makers for their daughters’ health, men may hold the final say and power in operationalizing that decision. In Indonesia, several mothers expressed a desire for greater involvement from fathers, reflecting a perceived gap in shared responsibility. In contrast, in Nigeria, fathers and male community or religious leaders often play a more overt and authoritative role in decisions concerning girls’ health. Some Nigerian mothers described scenarios in which men held the final authority over health matters, and women who acted independently risked social criticism—highlighting the persistent influence of traditional gender expectations. These findings illustrate the complex and context-specific nature of gender norms, particularly as they relate to intra-household and health-related power dynamics (48,49). This study, in which girls and women voiced a desire for greater involvement from fathers and male figures, highlights the importance of addressing gender norms and underscores the need to actively engage male decision-makers in HPV program planning. Recent studies have increasingly recognized that men play influential roles as decision-makers, household heads, and community leaders—and that their support can significantly influence vaccine uptake (50–53). Developing targeted outreach strategies to engage men as supportive partners in their daughters’ health and HPV vaccination efforts is essential (54). In Indonesia, this could include father-focused education sessions at schools and community events. In Nigeria, outreach through mosques, churches, and traditional gatherings may be more effective in increasing fathers’ awareness and support for vaccination.

Gender norms and expectations shape not only girls’ and their caregivers’ attitudes and access to the vaccine but can influence the effectiveness of vaccination campaigns themselves. This study highlights how social stigma and gender norms in Indonesia discourage girls from actively seeking information. In Nigeria, concerns about vaccine side effects and persistent myths about infertility remain major barriers, especially among unvaccinated girls and their caregivers. To effectively address these gender-related challenges, vaccination campaigns should consider the unique ways in which both girls and caregivers navigate health systems (55). For instance, stigma in Indonesia and fears of infertility in Nigeria require context-specific, culturally relevant messaging. Approaches like HCD offer value by engaging girls and their caregivers directly, ensuring that solutions are rooted in local realities and shaped by the voices of those most impacted (36). For HPV program planning specifically, HCD approaches can potentially improve uptake and sustainability (35,37,56).

### Limited Awareness and Information Gaps

Our findings reveal that both adolescent girls and their caregivers lack sufficient, consistent, and accurate information about HPV vaccination. Misinformation remains a significant barrier to uptake. While this information gap has been well-documented from the perspectives of parents and stakeholders, it has rarely been examined from the perspective of girls themselves (6–8,10,11,18,57). Our study makes clear that there is a widespread desire for more information; however, the content, messenger, and mode of delivery must be better aligned with the needs of the end users—namely, adolescent girls and their caregivers. Participants across both countries emphasized this point, with all proposing solutions centered on improving knowledge and awareness of the HPV vaccine. This underscores both the urgency of the issue and the need for a shift in communication approaches. In Nigeria, participants strongly recommended peer-to-peer education as an effective way to reach girls with accurate, relatable information. In Indonesia, participants stressed the need for more structured and engaging content, along with better training for teachers to deliver key messages effectively, as teachers were considered the most trusted source. The literature highlights that strengthening communication strategies to address mothers’ perceptions of HPV can significantly improve vaccine uptake among both mothers and their daughters (58–60). Furthermore, enhancing communication about the HPV vaccine may empower young people to take a more active role in their health care decisions, ultimately contributing to higher vaccination coverage. These insights highlight the critical value of involving end users in the planning and design of HPV communication strategies (37).

Ultimately, the challenge is not only about what messages are delivered but also who delivers them and how (61,62). Our findings suggest that current messengers are not always reaching or resonating with their intended audiences. Greater alignment between messenger and audience is needed to ensure communication efforts are trusted, accessible, and impactful. Specifically, strengthening vaccine education using trusted messengers (such as teachers, community leaders, and health workers) who deliver accurate, culturally appropriate information tailored to diverse audience needs is strongly recommended (54,56).

### Community Influence and Social Norms

This study highlights important cross-country differences in how social norms and decision-making structures influence HPV vaccination uptake in Indonesia and Nigeria. These findings underscore the importance of tailoring vaccination strategies to the local sociocultural context (63–65). In Indonesia, decision-making was primarily individualized and centered within the household. Mothers and daughters were often the primary decision-makers, and concerns about vaccination reflected personal or family-level considerations. This aligns with a systematic review on health-seeking behavior in Indonesia, which found that individualized circumstances were more influential than social factors (66). This suggests that in Indonesia, program strategies should focus on empowering households—particularly mothers—as key agents of change. Campaigns may be most effective when they emphasize the benefits of HPV vaccination for a girl’s health, future well-being, and academic or career success, using messaging that resonates at the family level.

In contrast, in Nigeria, decision-making is more collective and embedded within broader community dynamics. Religious and traditional leaders, along with peer networks, serve as influential gatekeepers. Drawing on the socio-ecological model, health choices are not simply personal— they are communal decisions, shaped by shared beliefs and group trust (67–69). Thus, HPV programs in Nigeria should prioritize community-based approaches, leveraging respected community figures to champion vaccination, combat misinformation, and foster a culture of acceptance. Establishing peer-to-peer networks can also help reinforce accurate knowledge and increase collective confidence in the vaccine (70,71).

These contrasts highlight a broader implication: understanding the social ecosystem—who makes decisions, who influences them, and what values they are anchored in—is essential for developing effective, context-sensitive strategies that can lead to higher and more equitable HPV vaccination coverage. In more family-oriented settings, interventions should speak directly to the family unit; in communal settings, they must ripple outward through social structures and trusted voices. Recognizing who the reference groups are in each setting—whether it’s a mother, a religious leader, or a peer—allows programs to align their approaches and messages with local values and decision-making pathways.

### Service Delivery: In-School vs. Out-of-School Girls

Effective HPV vaccination requires not only demand generation but also reliable and equitable service delivery. Our findings point to important differences in how service delivery challenges affect vaccine uptake in Indonesia and Nigeria, with distinct implications for reaching both in-school and out-of-school girls.

In Indonesia, the school-based model has generally provided a consistent platform for vaccine delivery. However, this approach leaves significant gaps for out-of-school girls, who often face informational rather than logistical barriers (72). Moreover, the current model does not adequately create a welcoming environment or recognize that girls may have questions and need support during the vaccination process. Opportunities exist to improve the school experience by reducing wait times and fostering support through storytelling activities and more engaging interactions with teachers and health workers.

Service delivery challenges in Nigeria are more systemic, multifaceted, and align with the existing literature on HPV barriers (73). Even when girls and caregivers were informed and motivated to seek vaccination, they frequently encountered structural barriers: long wait times, vaccine stockouts, transportation difficulties, and insufficient infrastructure all limited their ability to access vaccines (1,73–75). These barriers particularly affect out-of-school girls, for whom health services may be even more out of reach due to geographic, financial, or social constraints (72).

These findings carry several important implications for program design. First, school-based vaccination alone is insufficient to achieve equitable coverage (76). Out-of-school girls—who are often among the most vulnerable—require alternative and flexible delivery mechanisms. Mobile clinics, house-to-house outreach, and partnerships with community health centers and religious institutions can help bridge this gap (72,76). In Indonesia, mobile outreach combined with trusted local institutions may extend the reach of services beyond the classroom. In Nigeria, these approaches are even more critical, given the breadth of logistical challenges that impede access.

Second, there is an urgent need to strengthen health system capacity and planning. In Nigeria, this includes improving supply chain management, investing in health infrastructure, and training frontline workers to deliver vaccines more efficiently and sensitively. In Indonesia, improvements in communication, standardization of procedures, and support and training for health workers and teachers can improve the quality and consistency of school-based delivery.

Ultimately, the needs of in-school and out-of-school girls differ not just in terms of access points, but also in the types of support they require. A flexible, equity-focused service delivery model that recognizes these differences is essential to improving HPV vaccination coverage and ensuring no girl is left behind.

## Conclusion

This study is one of the few that places girls at the forefront of HPV vaccine program planning, highlighting the value of designing interventions around the needs and lived experiences of the end user. By centering girls, HPV vaccine programs can better tailor strategies that not only improve uptake but also enhance long-term sustainability. Importantly, the findings point to the potential of peer-to-peer education as a powerful strategy, leveraging girls’ own social networks to increase trust, correct misinformation, and strengthen vaccine confidence.

The findings underscore the critical need for context-specific, gender-responsive approaches to increase HPV vaccination in both Indonesia and Nigeria. In both settings, persistent misinformation and limited awareness call for targeted communication strategies delivered by trusted messengers. Reaching out-of-school girls also demands flexible, equity-focused service delivery models that extend beyond school-based platforms.

Ultimately, a one-size-fits-all approach is inadequate—HPV programs must be grounded in local decision-making structures, gender norms, and access barriers to ensure broader and more equitable vaccine coverage.

## Data Availability

De-identified qualitative data available upon request.

## Acknowledgement

We want to acknowledge all the data collectors in both Indonesia and Nigeria who made the collection of this research possible. This material was produced by the HPV Vaccine Acceleration Program Partners Initiative (HAPPI) Consortium and may be used freely. The HAPPI Consortium, in collaboration with our esteemed partners, is managed by JSI together with Clinton Health Access Initiative (CHAI), the International Vaccine Access Center (IVAC) at the Johns Hopkins Bloomberg School of Public Health, Jhpiego, and PATH. This research and the HAPPI consortium are funded by the Gates Foundation under grant number INV-057603.

## Notes

### Competing Interest Statement

The authors have declared no competing interest.

### Funding Statement

This research was funded by the Gates Foundation under the grant number INV-057603.

### Author Declarations

The study was approved by JSI Institutional Review Board (IRB) under the IRB reference (#24-37). The JSI IRB is registered with the Office of Human Research Protections (OHRP) as IRB00009069. JSI has a Federal-wide Assurance (FWA) on file [FWA00000218] and is a recognized IRB organization [IORG0007551]. Local ethics approval was received in both Indonesia and Nigeria. Written consent was obtained by all study participants.

### Summary of Updates

No major difference, slight update to methods.

